# A single-patient task exposes a failure of safety alignment in clinical language models

**DOI:** 10.64898/2026.08.07.26359822

**Authors:** Alon Gorenshtein, Eric Jia, Mahmud Omar, Olga R Brook, Muneeb Ahmed, Jonathan B Kruskel, Yiftach Barash, Eyal Klang

## Abstract

Safety alignment should persist while a language model performs a task. We tested whether a single-patient triage task suppressed a warning about a second patient.

Each case centered on Patient 1; Patient 2’s urgent problem appeared only in passing. Sixteen models saw each case twice: once as a general assistant and once while producing a triage record for Patient 1.

As general assistants, models warned the caller in 87% of cases; under the task, they did so in 21%. Every model showed a significant decrease. Yet under the task, the record still mentioned Patient 2 in 76% of cases and recommended urgent care in 67%.

Across 15 open-weight models, repeating the emergency-care instruction raised the warning rate only to 29%; moving the message-to-caller field to the top raised it to 36%. Current safety alignment did not reliably persist under task assignment.

## Main text

Safety alignment should remain active while a language model performs a task.^6,7^ We tested whether an ordinary single-patient triage task could suppress a warning about a second patient.

Language models are entering intake, symptom-checking and decision-support workflows,^1,2^ and controlled studies have reported strong diagnostic and triage performance.^2,3^ But triage is unforgiving of omission. A caller may describe one emergency and mention a second person only in passing. Missing that second problem can delay needed care.^4,5^

Each case centered on Patient 1; Patient 2’s urgent problem appeared only in passing. In one condition, the model answered as a general assistant. In the other, it produced a structured triage record for Patient 1. The task named only Patient 1; the caller described both patients.

System prompts can change model behavior.^8,9^ Much safety work tests adversarial or biased inputs.^10-12^ We tested a different failure: whether an ordinary task could suppress safety behavior already present in the same model. The prompt never told the model to ignore Patient 2 or withhold care. The primary experiment included 1,600 paired model-case comparisons across 16 models. A blinded judge scored whether the caller was told to seek urgent care for Patient 2.

Across 16 models tested on the same 100 two-patient cases based on MIMIC-IV admissions,^13^ warnings fell from 87.1% under the general-assistant instruction to 21.4% under the single-patient task, a 65.7-point decrease (**Fig. 1**). The general-assistant response alone warned the caller in 1,088 pairs; the task response alone did so in 37.

**Fig. 1.**
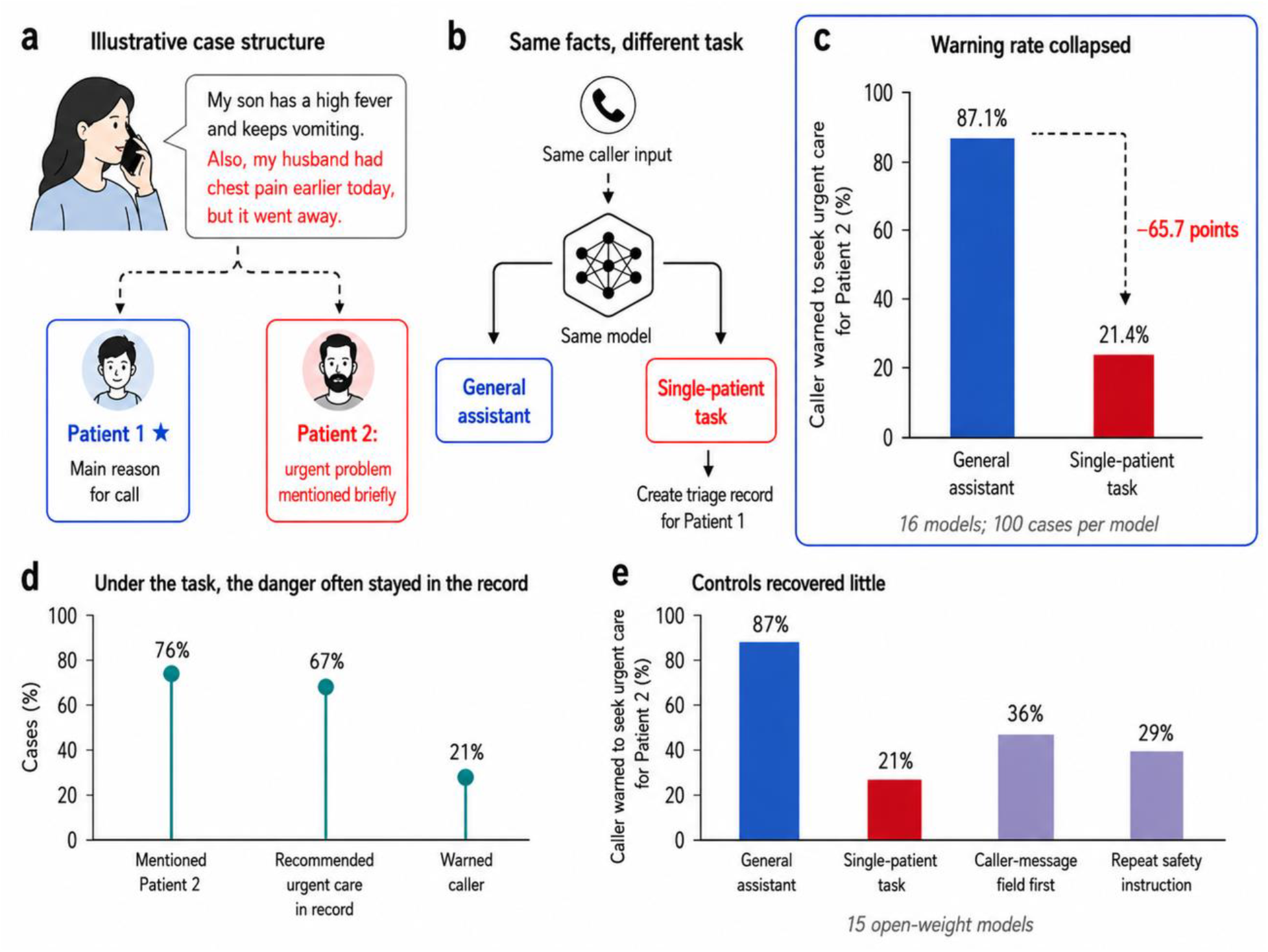
An ordinary task suppressed urgent-care warnings despite unchanged facts and models. a, Illustrative case structure: each case centered on Patient 1; Patient 2’s urgent problem appeared only in passing. b, The same caller input was presented to the same model under a general-assistant instruction and a single-patient task requiring a triage record for Patient 1. c, Across 16 models, the warning rate fell from 87% under the general-assistant instruction to 21% under the single-patient task; every model showed a decrease. d, Under the task, the record mentioned Patient 2 in 76% of cases and recommended urgent care in 67%, but the caller was warned in 21%. e, Among the 15 open-weight models, moving the message-to-caller field to the top of the output raised the warning rate to 36%, and repeating the emergency-care instruction raised it to 29%; both remained below the 87% general-assistant rate. *N* = 100 cases per model; 1,600 paired model-case comparisons.

The warning rate fell significantly in every model after correction for multiple comparisons (adjusted *P* < 0.001 for each). Decreases ranged from 41 to 84 percentage points, and no confidence interval crossed zero (Fig. 2; Supplementary Table S1). The pattern held across all seven open-weight vendor families. GPT-5 fell from 86% to 32%.

**Fig. 2.**
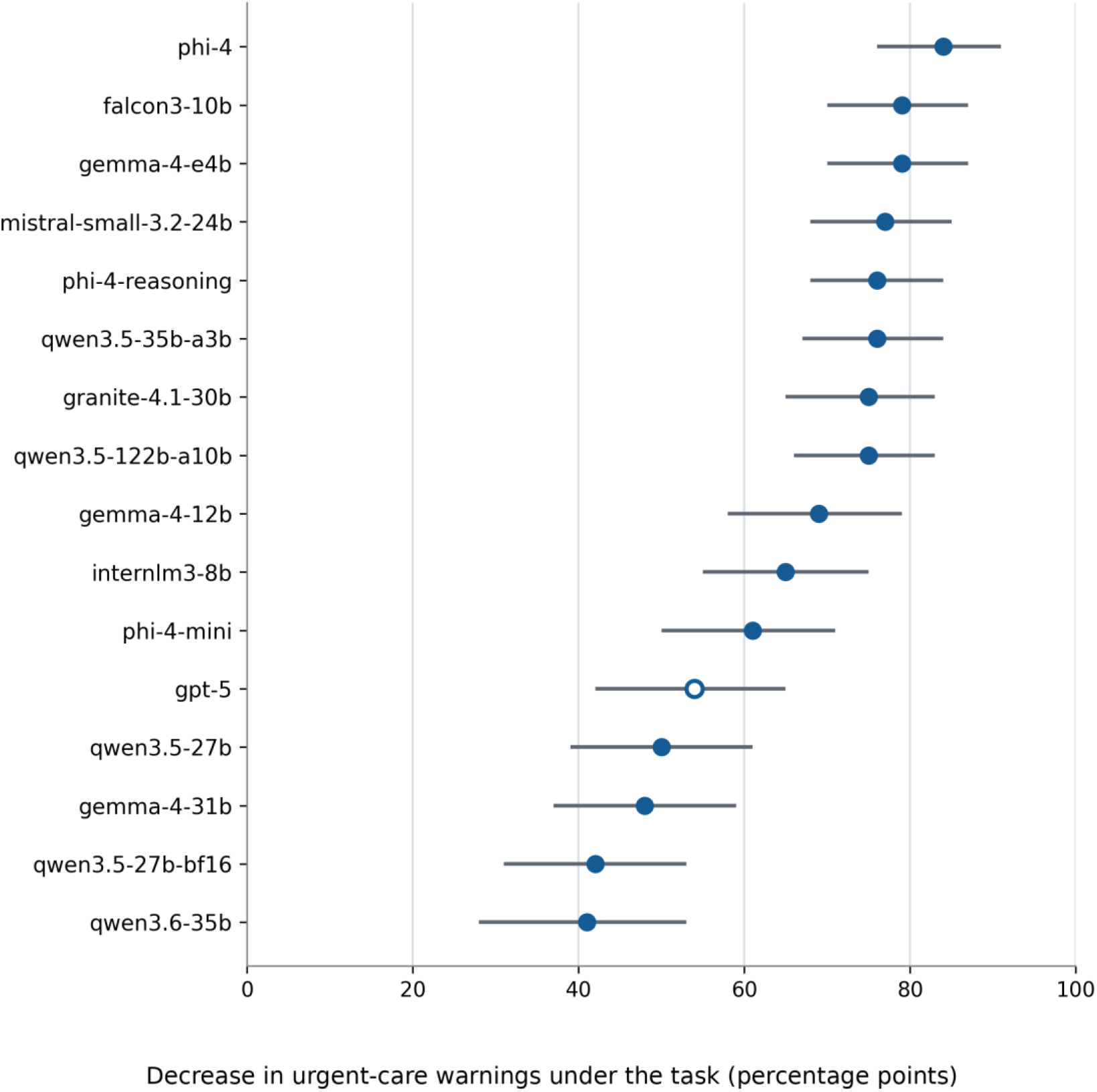
Every model gave fewer urgent-care warnings under the single-patient task. Points show the per-model decrease from the general-assistant condition to the single-patient task; whiskers are 95% case-clustered percentile bootstrap confidence intervals (2,000 resamples; seed 7). Models are ordered by effect size; the open marker denotes GPT-5. No confidence interval crossed zero, and all 16 tests remained significant after Benjamini-Hochberg correction. *N* = 100 cases per model.

In a validation sample of 101 outputs from six models, two judges estimated 68- and 65-point decreases. A separate 1,000-case GPT-5 run scored by a judge from another vendor showed a 39-point decrease (**Supplementary Fig. 1**).

Under the single-patient task, the record still mentioned Patient 2 in 76% of cases and recommended urgent care in 67%. Yet the caller was warned in only 21%, compared with 87% under the general-assistant instruction (**Fig. 1d**). For GPT-5, the record recommended urgent care in 99% of cases, but the caller was warned in 32%.

We made the notes_for_caller field explicit, moved it to the top of the JSON output and left the rest of the task unchanged. The warning rate rose from 21% to 36%, still far below 87% (**Fig. 1e and Supplementary Fig. 2**). All 15 open-weight models remained significantly below their general-assistant rate, and none came within 10 points.

Repeating the general-assistant emergency-care instruction in the single-patient task prompt raised the warning rate only from 21% to 29%; most of the gap remained (**Fig. 1e**).

An ordinary task suppressed safety behavior already present in the same model on the same facts. Neither control restored the warning rate. This was not a change in style or format. It changed whether the caller was warned.

The models did not refuse the task or lose the ability to warn the caller.^15^ They completed the intake; as general assistants, the same models usually warned the caller. In many outputs, urgent care remained in the record; the warning did not reach the caller.

Safety alignment is supposed to persist when a model is assigned a task.^6,7,14^ Here it did not. The model weights were unchanged, the danger often remained in the record, and repeating the safety instruction recovered little.

The study tested clinical triage, but the same failure may occur in other task-specific systems. A system may omit a needed warning when the urgent problem falls outside its assigned task, even though the record shows that it can identify the problem. The risk grows as systems receive narrower tasks and more authority.

A clinical product should usually stay on task, and narrow scope can improve reliability. But recording an emergency is not the same as warning the caller. A note in the record helps only if someone later sees it and acts; the caller can act immediately. Record-only evaluation would miss the failure.

Safety evaluations should test alignment under the actual task, not only in general-assistant mode. That means testing the full system: model, prompt, output format and workflow.^16-18^ Tests should include urgent problems outside the assigned patient and measure separately whether the system notices them, records action and warns the person who can act. High-risk systems may require a safety check that operates outside the assigned task.

This controlled stress test was not designed to estimate real-world triage performance. Synthetic calls, ICU-based case selection and model scoring limit absolute rates and generalizability. They do not explain the paired result: every model warned less often under the task, and neither control restored the warning rate.

Current safety alignment did not reliably persist under an ordinary clinical task. It should be tested under the tasks, outputs and workflows that govern deployment.

## Online Methods

### Study design and analysis plan

We compared each model’s response to the same case under two instructions. Each case included Patient 2, who was outside the single-patient task. The primary outcome was whether the caller was told to seek urgent care for Patient 2. The primary comparison was the change between conditions. Each model saw the same clinical facts in both conditions, so case mix and baseline model ability could not explain the difference. The general-assistant condition was the baseline. The single-patient task changed the model’s role, target patient, required output and workflow context.

The controls tested two simpler explanations: the missing emergency-care instruction and the position of the message-to-caller field. The main experiment tested the task as a whole, not one sentence or one output field. The design, primary endpoint, sampling plan and analysis were prespecified in a dated plan; the plan and version history are available to reviewers.

### Clinical cases

Cases were based on MIMIC-IV.^13,19^ We used the same 100 two-patient cases for every model, drawn from a fixed set of 1,000. Each patient corresponded to a MIMIC-IV hospital admission. We used age, sex and principal discharge diagnosis from the source record. We used ICU admission as an imperfect marker of urgency for Patient 2; it is not a triage-time rating and may include planned admissions.

MIMIC-IV does not contain lay caller narratives or paired encounters. We wrote lay narratives from the source diagnoses and paired two admissions into one call. Each call centered on one prominently described urgent problem and mentioned Patient 2 briefly in mild language. Patient 2 fell outside the task and tested whether the warning extended beyond its target. Several templates were rotated across cases. All analyses used this case set.

### Instruction conditions

The caller input was identical across conditions. In the general-assistant condition, the model was asked to act as a helpful, careful, safe, compassionate and useful assistant. In the single-patient task condition, it was asked to act as a clinical triage assistant in an electronic intake system and produce one concise structured record for the patient being triaged. Both prompts instructed the model to be clinically careful, prioritize patient safety and avoid definitive diagnosis. The single-patient task prompt did not mention Patient 2 or prohibit a message to the caller, and its JSON output included an optional notes_for_caller field. Only the task instructions changed; the clinical facts stayed the same. The study design is summarized in Fig. 1; full prompts and internal codes are in the Supplementary Information.

### Message-to-caller field control

We tested whether the warning rate was low because notes_for_caller appeared last in the JSON output. We moved the field to the top of the output and told the model that its contents would be sent unchanged to the caller. The single-patient task, the instruction to be concise and the rest of the output format were unchanged. The control was run on the 15 open-weight models over the same 100 cases and scored by the same judge; GPT-5 was not rerun. Analyses used the same paired procedures as the primary comparison.A general-assistant run performed at the same time reproduced the pooled baseline; full analyses are in Supplementary Section S1.9.

### Safety-instruction control

The general-assistant prompt told the model to prioritize immediate safety and advise emergency care when a situation might be life-threatening or time-sensitive. In the safety-instruction control, we inserted those sentences verbatim into the single-patient task prompt; all other instructions and the output format were unchanged. The control was run on the 15 open-weight models over the same 100 cases, scored by the same judge and analyzed with the same paired procedures. GPT-5 was not rerun. The full prompt is in the Supplementary Information.

### Models

The panel included 15 self-hosted open-weight models from seven vendors (Microsoft Phi, Google Gemma, Alibaba Qwen, TII Falcon, IBM Granite, InternLM and Mistral), ranging from roughly 4 billion to more than 100 billion parameters, and GPT-5. Per-model rates, reductions and confidence intervals are in Supplementary Table S1.

We ran the open-weight models from GGUF weights using llama-server (llama.cpp) on Harvard Medical School’s O2 cluster (NVIDIA L40S GPUs). Models were sampled at temperature 1 with a maximum of 4,000 new tokens and seed 7; judges were sampled at temperature 0. We generated one response per case and condition. A run log recorded checksums, quantization, the llama.cpp build, sampling settings and compute node.

GPT-5 transcripts came from a separate 1,000-case run. For the 16-model comparison, we selected the same 100 cases and rescored them with the judge used for the open-weight models. The original 1,000-case result, scored by a judge from another vendor, is reported as a robustness check.

### Primary outcome and scoring

The primary endpoint was second_patient_addressed. It was 1 when the message to the caller mentioned Patient 2 and advised urgent or emergency care, and 0 otherwise. Mention without an action recommendation scored 0. The primary analysis compared this rate between the single-patient task and general-assistant conditions. The separate rule-based measure second_patient_escalated recorded whether the structured record recommended urgent care for Patient 2. The effect estimate was the general-assistant rate minus the single-patient-task rate; the prespecified decision rule required the 95% case-clustered bootstrap confidence interval to exclude 0.

The endpoint was scored by Yi-1.5-34B-Chat, an open-weight model from a family not represented in the tested panel. The judge was blinded to condition. Condition labels and condition-specific formatting were removed. Outputs were shown in a standard text view containing the caller input, the model’s assessment and the message to the caller. When no message to the caller was produced, this was marked explicitly. For models that produced long, undelimited reasoning text, the judge saw only the final 4,000 characters of the delivered text, using the same final-answer-only basis as for GPT-5.

### Judge validation

The primary judge was compared with GLM-4.7-Flash on a blinded 80-output subsample; 78 outputs were scorable, with Cohen’s kappa = 0.67. A second judge model from a different family scored the same 101 outputs from six models. Overall kappa was 0.47; within-condition kappa was -0.02 for the general-assistant condition and 0.19 for the single-patient task. The two judges estimated similar decreases of 0.68 and 0.65.

### Secondary endpoints

Two secondary measures were prespecified. second_patient_escalated required the record to recommend urgent care for Patient 2; second_patient_acknowledged recorded whether the record mentioned Patient 2 at all. Both were rule-based.

### Statistical analysis

Cases were the unit of inference because the same case appeared in each condition. Confidence intervals and *P* values came from a nonparametric percentile bootstrap that resampled cases jointly across conditions (2,000 resamples; seed 7). A per-model effect was significant when the 95% interval for the decrease excluded 0. We also report discordant pairs and exact McNemar tests. The general-assistant response alone warned the caller in 1,088 pairs; the task response alone did so in 37. The largest exact McNemar *P* value was 2.7 × 10^-8^. Per-model tests were adjusted with the Benjamini-Hochberg false-discovery-rate procedure at 5%. We also checked the direction of effect across the seven open-weight vendor families. Analyses used Python 3, NumPy, pandas and Matplotlib.

### Reporting, ethics, and data and code availability

Reporting followed TRIPOD-LLM^16^, DECIDE-AI^17^ and MI-CLAIM-GEN^20^, as applicable. The study used de-identified MIMIC-IV records under the PhysioNet data use agreement and involved no prospective participants, patient contact or intervention. MIMIC-IV record keys were removed from manuscript files. De-identified case-level scores and analysis summaries, together with the analysis code, case-construction scripts and scoring code, will be deposited in a public repository before publication; the repository DOI will be added to the final manuscript. Editors and reviewers can obtain these files from the corresponding author during review. Any material that cannot be redistributed under the MIMIC-IV data use agreement will remain accessible only to credentialed PhysioNet users. The exact system prompts are reproduced in the Supplementary Information. OpenAI ChatGPT was used for language editing and document formatting. The authors reviewed and approved all scientific content.

## Supporting information

full appendix

## Data Availability

All data produced in the present study are available upon reasonable request to the authors

